# Sub-weekly cycle uncovers the hidden link of atmospheric pollution to Kawasaki Disease

**DOI:** 10.1101/2020.06.04.20122325

**Authors:** X Rodó, A Navarro-Gallinad, T Kojima, J Ballester, S Borràs

**Affiliations:** ICREA, Barcelona, Catalonia, Spain.; Climate and Health (CLIMA) Program, ISGlobal, Barcelona, Catalonia, Spain.; ADAPT Centre, Trinity College Dublin, Dublin, Ireland.; Faculty of Advanced Science and Technology, Kumamoto University, Kumamoto, Japan

## Abstract

Anthropogenic pollution has frequently been linked to myriad human ailments despite clear mechanistic links are yet lacking, a fact that severely downgraded its actual relevance. Now a prominent unnoticed sub-weekly cycle (SWC) of 3.5 days is uncovered in the long-term epidemiological records of Kawasaki disease (KD) in Japan, a mysterious vasculitis of yet unknown origin. After ruling out the effect of reporting biases, the analysis of Light Detection and Ranging (LIDAR) atmospheric profiles further confirms that this variability is linked to atmospheric particles with an aerodynamic diameter less than 1 µm. SWC accounts for 20% of the variance in KD and its contribution is stable throughout the entire epidemiological record dating back to 1970, both at the prefecture level and for entire Japan. KD maxima in 2010-2016 always occur in full synchrony with LIDAR particle arrival in diverse locations such as Tokyo, Toyama and Tsukuba as well as for the entire of Japan. Rapid intrusion of aerosols from heights up to 6km to the surface is observed with KD admissions co-varying with their metal chemical composition. While regional intensity of winds has not changed in the interval 1979-2015, our study instead points for the first time to increased anthropogenic pollution as a necessary co-factor in the occurrence of KD and sets the field to associate other similar human vasculitis.

## Introduction

Causes conducive to Kawasaki disease (KD) syndrome are yet unknown, despite the intensive research on different fronts (genetic, immunological, environmental, epidemiological, etc., 1,2). Alleged drivers include a myriad of potential factors under different hypothesis around the innate and adaptive immune responses observed in KD (e.g. bacteria, fungi, viruses and toxins, 3–8) but no conclusive result has unequivocally solved this long-standing puzzle. KD is an acute, self-limited vasculitis that occurs in children of all ages. On the etiology, the leading paradigm is that a yet unidentified agent enters through the upper respiratory tract and causes a dramatic immunologic response in certain genetically predisposed children (9,10). Coronary artery aneurysms develop in 20–25% of cases, with this development being at times clinically silent and often misdiagnosed with other more benign rash/fever syndromes caused by viruses or bacterial toxins (11). It may lead years later, to sudden death or myocardial infarction (12). Seasonality of KD was first described for Japan(13) and then worldwide (14). Increases in KD cases in winter in locations at both sides of the North Pacific Ocean were seen to occur associated with the seasonal enhancement of low- and high-tropospheric wind currents from the Asian continent (15). The ulterior identification of a fungal pathogen being predominant in the aircraft profiles in Japan made on air masses over the planetary boundary layer (PBL) at times of KD maxima, led to the hypothesis that KD could be caused by a pathogen or environmental trigger carried out by winds blowing from the Asian continent (8,15–17).

The long rich record of epidemiological incidences of KD in Japan going back to the seventies enables a very detailed and unprecedented statistical approximation (8,14,15,18). Now detailed analyses of daily admissions of KD in epidemiological records for the entire of Japan and for 9 regions grouping prefectures, clearly uncover the presence of a prominent sub-weekly cycle (SWC) along with the already known strong seasonality as the dominant variability components (Fig. 1A, B). Singular Spectrum Analysis (SSA) decomposition (M = 100; see Methods and 19,20) applied to these datasets separated among significant cyclical components that accounted for most of the variability in the time series. Figure 1 displays for the historical daily epidemiological records of KD aggregated for entire Japan, both the seasonal evolution (Fig. 1A) and a prominent SWC (Fig. 1B). An AR spectrum with a backward-forward singular value decomposition algorithm was computed for KD (Fig. 1C) to further determine the sources of spectral variability and assess the nature of the weekly oscillations. First two Reconstructed Components (RC) recovered seasonality (RC12, 70% of the variability, *p< 10^−5^*; Fig. 1A), and a previously unnoticed SWC showed up strongly that accounted altogether for a 20% of the variance. This SWC was composed by two clearly distinguishable sub-periods (Fig. 1C), with the fundamental one being that of 3.5 days (d) (SWC1 >10.6%; *p< 10^−5^*), and harmonic at 2.33d (SWC2 = 9.2%; *p< 10^−5^*, see methods).

**Figure 1:**
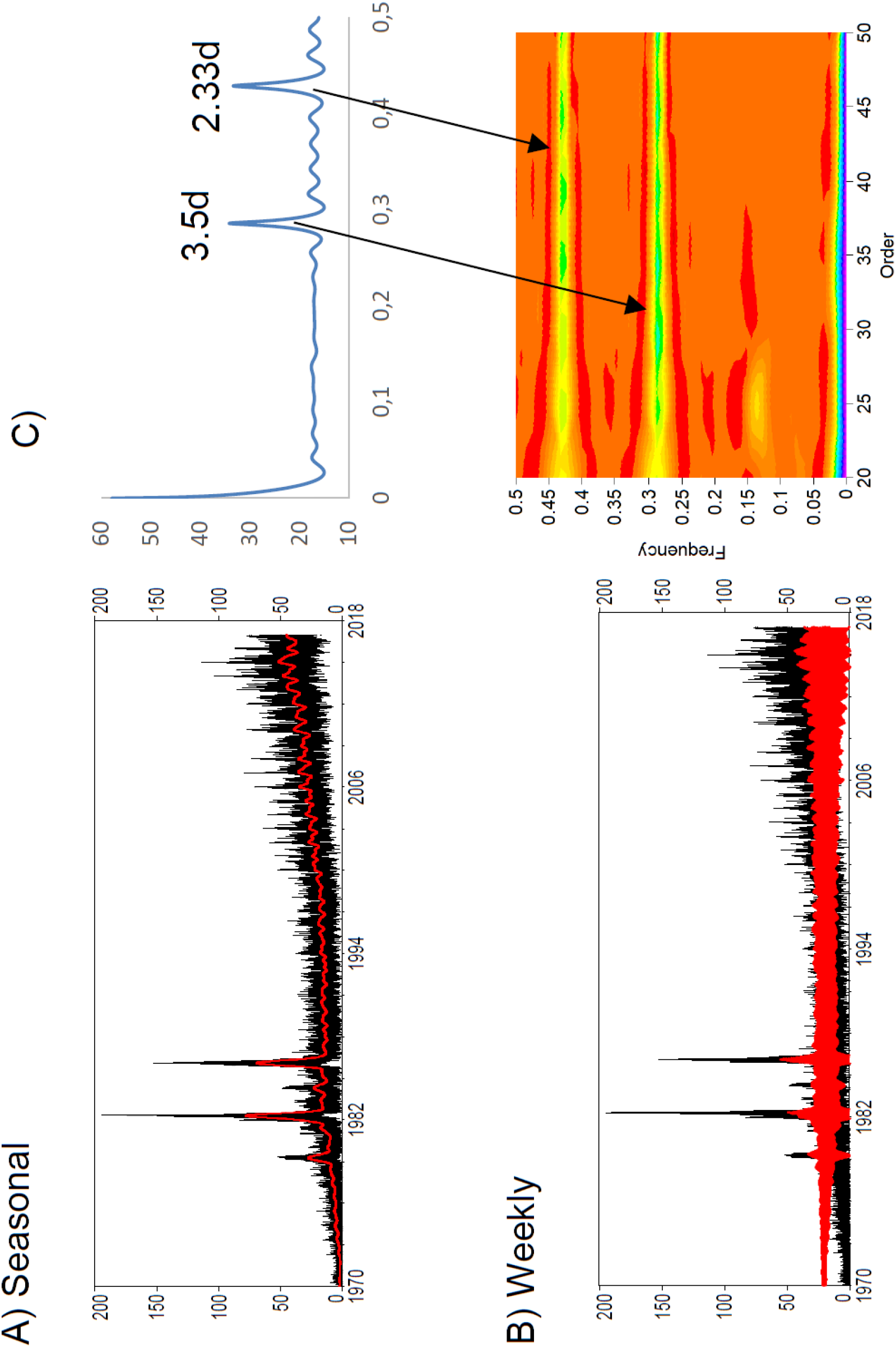
Long-term evolution of Kawasaki disease (KD) incidence for entire Japan (1970-2016) and weekly cycle contribution. A) Aggregated KD incidence for all of Japan (black) and reconstructed variability at the seasonal to interannual timescales (red, corresponding to RC12 of the SSA decomposition; see Methods for spectral decomposition and reconstruction (*19*). B) Same as A) but for the weekly cycle variability, decomposed into individual cycles in C). Top panel: AR Spectrum of the KD time series with FB (forward-backward) singular value decomposition algorithms and bottom panel: results for individual sub-weekly frequencies obtained (namely 3.5d and 2.33d, respectively), and exploration of different orders for stability of results.

Existence of (seven days) weekly cycles in human diseases -or in clinical data- has been attributed to a surrogate of the weekday-weekend dynamics in hospital admissions in poor health care locations with reduced staffing levels or less experienced staff (21–23), or to lower number of admissions for mild diseases or even to biases in reporting (e.g. admissions being assigned to the following Monday or labor day, 24). For instance, difference in medication outcomes and even death can occur due to lower quality or activity of medical care, or even delayed initiation of treatments in the case of severe diseases (25,26). Lower admission rates also occur over the weekend for asthma and mild diseases (27) and in prevalence surveys of nosocomial infections, due to the effects of air pollution on hospital admissions and mortality (28). But most previous studies on weekday variability have focused on acute admissions, such as an increased risk of myocardial infarction, ischemia, sudden death, and cardiac arrests occurring on Mondays (29–33). Conversely, weekend admission in Japan was associated with increased mortality in patients with severe community-acquired pneumonia (23) and also with increased myocardial infarction in women (34). The association of mortality with day and time of admission has been tackled by many studies, with considerable heterogeneity in the results (25).

It is well-known that no natural weekly cycles exist, being the weekly period a cycle with a unique anthropogenic fingerprint (35–37). There are natural daily circadian rhythms, seasonal or sub-seasonal cycles and long term interannual changes (e.g. El Niño-Southern Oscillation, decadal variability…). Human activity in industrialized countries largely follows a 7d cycle, where fossil fuel combustion is expected to be reduced during weekends (38–41). This weekend effect is well known from local, ground-based measurements, and may even translate into a small temperature signature, associated to a reduction in transportation (42). Nitrogen oxides (NO+NO_2_=NO_x_ and reservoir species) are important trace gases in the troposphere with impact on human health, atmospheric chemistry and climate (e.g. due to ozone production, 43; and their influence on the OH concentration, 44). Some studies showed the weekly cycles of tropospheric NO_2_ for different regions of the world (42), with a clear minimum in the US, Europe and Japan and even with 35% lower NO_2_ levels on Sundays than on working days in Germany (45).

In the current study, we first separate KD variability and address the origins of the SWC components from the long epidemiological records of KD in Japan. After carefully discarding the influence of a potential weekend bias, we move towards tracing the 3.5d SWC in KD to temporally co-vary with the same variability existing in atmospheric particulate matter (PM_x_) and wind dynamics over Japan. Atmospheric PM_x_ composition and movement are then tracked by means of the Asian dust and aerosol LIDAR observation network (AD-Net) (46,47). LIDAR profiles are also scrutinized to search whether wind intrusions from the mid-troposphere are associated to both aerosol peaks and KD peak maxima seasons. We further describe the time taken by these air intrusions to affect and originate recognizable KD increases. Finally, we investigate the potential relationship of KD incidence to the PM_x_ chemical composition, specially to airborne metals.

To determine the base frequencies in daily KD records, we conducted a two-way Scale-Dependent Correlation (TW-SDC) analysis (48–50) as it is specifically aimed at uncovering transitory associations between joint variability structures in time series (see methods). We used the daily series of KD in the recent interval for which we also have LIDAR atmospheric profiles (2010 in Fig. S1 and January 2010-December 2016 in Fig. S2), and a synthetic time series of 3.5d (SWC1), 2.33d (SWC2) and 7d (SWC3) periods (see Methods). Fig. S1 displays the TW-SDC for SWC1(a,d), SWC2 (b,e) and SWC3 (c,f) at different window sizes, s (s = 6d in a,b,c and s = 22d in d,e,f) for the year 2010. The prominent role of the 3.5d cycle (SWC1) stands out clearly, both in terms of correlation magnitude as well as of presence throughout 2010–2016 (a *vs* c, d *vs* f and Fig. S2). In addition, a very faint SWC3 (7d) essentially derived from the 3.5d cycle, correlates instead to periods with very low KD incidence (Fig. S1c and f). A similar analysis with an intermediate s of 10 to maximize a potential 7d cycle applied to the entire 2010–2016 interval further reinforces instead the more dominant role of SWC1 in KD (Fig. S2). This way, two maxima and two minima clearly exist in a week and the SWC strongly shows up in all the large epidemics (Fig. 2A, for the largest KD epidemic, in 1982; Fig. S3 for 1979, 1982 and 1986). SWC1 exists throughout the entire KD epidemiological record since surveillance was initiated in the early 1970s in Japan (and up to the latest data available at the end of 2016, Fig. 2B). Concomitant capricious behavior at both minima and maxima during the largest epidemic is not consistent with a systematic reporting bias on Sundays-Mondays (8).

**Figure 2.**
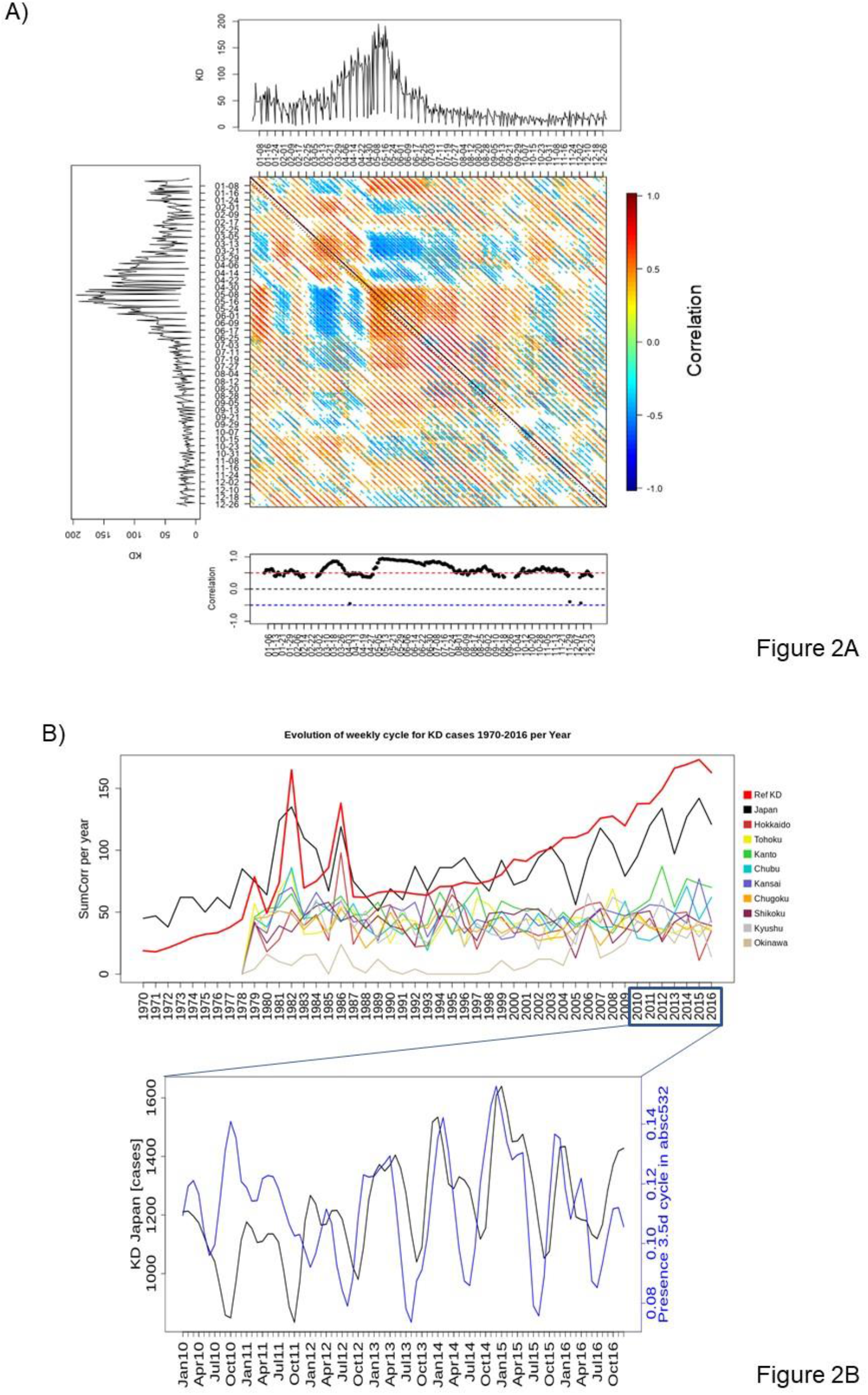
Weekly cycle in KD throughout the historical epidemiological record (1970-2016) A) Example of an SDC analysis applied to the daily KD epidemiological dataset aggregated for the entire Japan during the 1982 epidemic (see Fig. S3 for the three largest epidemics). Due to the dataset daily resolution, the SDC cannot resolve cycles shorter than 4 days, albeit the resulting 7d SWC clearly shows up as alternating diagonal stripes. Notice the absence of the periodicity only in short intervals. B) Presence of the SWC in the long-term temporal reference evolution of KD incidence in Japan (red, relative scale), for all Japan prefectures altogether (black) and for each of the nine main regions in Japan (colored). Y-axis displays the sum of days with positive correlations close to the diagonal per year at lag 7 obtained from a one-way SDC (OW-SDC) analysis of the KD time series at s = 7d and alpha = 0.05 (Y). In the X-axis the time interval is in years (X). Evolution follows the same behavior in the maxima and linear ascending trend after them. This last variable is set as a reference and it has been scaled according to the plot dimensions. C) Average presence of the SWC1 (3.5d) in the four absc532 LIDAR atmospheric layers and evolution of the KD monthly sum of cases. Values on the Y-axis are the weekly cycle mean presence magnitude along the four atmospheric layers (see Methods for an explanation of methods to quantify the presence of the weekly cycle).

To assess whether geographical coherence exists for this SWC1 period throughout Japan, a specific analysis tracing the presence of the 3.5d was conducted for the main 9 large aggregations of prefectures in Japan (Fig. 2B) (see Methods). As seen, contribution of this variability is roughly constant throughout the entire record (seen since 1977 when separate prefecture information is available).

The former stability in the SWC1 amplitude is true also for all groups of individual prefectures, irrespective of their population, with the exception of Okinawa for which there is a marked enhancement of the SWC in the last few years (Fig. 2B). Despite, as said above, no natural cycles are described that have a weekly or semi-weekly periodicities, we further checked whether bias-reporting could have generated those persistent periodicities. The latter was totally discarded as the SWC cycle amplifies towards the present concomitant with the appearance of coherent marked seasonality in the epidemiological time series (8,15). Fig. S3 shows the reconstructed SWC(top) for the three main epidemics in Japan (1979, 1982 and 1986), together with the individual contribution of the different RCs of the SWC, namely SWC1 (3.5d; second row), SWC2 (2.33d; third row). Daily OW SDC analysis of KD per prefecture at s = 7d was further conducted and strong positive correlations close to the diagonal counted and grouped annually for the entire of Japan (Fig. 2B, black line; see Methods). Results shows a clear enhancement of the SWC1 towards the present, a rising trend and also an intriguing 4-year oscillation, discussed elsewhere (8). Total yearly aggregated KD incidence for Japan (red line) is there shown for comparison (Fig. 2B), with a slightly steeper upward trend towards the present, as well as an absence of the formerly described 4-year period. This same 4-yr period can be seen in the bottom panel of Fig. 2B, computed with monthly data instead. The increase in the number of times air intrusions is associated with more KD events might be thought to be driven by changes (increases) in wind intensity. Instead, close analyses show that despite the mean gradient of winter (DJF) regional wind stress for the interval 1975–2015 in northeast Asia clearly points to a straight connection between NE Asia and Japan (8) (Fig. S4A), there were not regional changes in the trends of wind stress nor in zonal and meridional wind speed during winter occurred (Fig. S4B). Instead warming indicates rises in northeast Asia of more than one degree Celsius on average from the 2000s with respect to the seventies. Therefore, mechanisms must point either or both to a gradual optimization in the direction of source winds -as indicated by the recent phase-locking of the seasonal variability in both KD and winds(15)-, or to an increase in the aerosol load due to enhanced anthropogenic activity in the source region.

To test for this possibility, we traced the SWC1 in KD, and its relation to anthropogenic activity (e.g. pollution and aerosols). The latter was investigated by means of LIDAR measurements, available from 2010 onwards. Fig. 3A displays the relation from 2010 to 2016 of KD monthly total cases in Japan (black) and a surrogate variable tracing the presence of SWC1 in particles in the Tokyo air column (SWC1-PM_1_ for absc532, blue). Absc532 stands for 532nm attenuated backscatter coefficient from LIDAR measurements averaged at both the surface (Fig. 3A, 4A) and at different height levels for Tokyo, Toyama and Tsukuba (Fig. 4B). Attenuated backscattering ratio between 1064nm and 532nm (absc1064/absc532) has been described to be an excellent analog for sizes of aerosol particles (51,52), yielding here values well below 1 (not shown) and therefore clearly denoting dominance of particles of size below 1µm (Fig. S5). Amount of PM_1_ aerosols clearly grows during events associated to KD maxima (Fig. S5A). Fig. 3A displays a striking monthly (weekly in Fig. 4A) positive coevolution between the SWC1-PM_1_ cycle (blue) and KD incidence (black), with the presence of the 3.5d cycle shortly anticipating KD (Fig. 3B). Synchronous annual cycles show up in both variables, with coincident maxima in winter and minima in summer, as expected. This large seasonal coherence among wind and SWC1-PM1 further reinforces the central role of aerosols in this disease.

**Figure 3.**
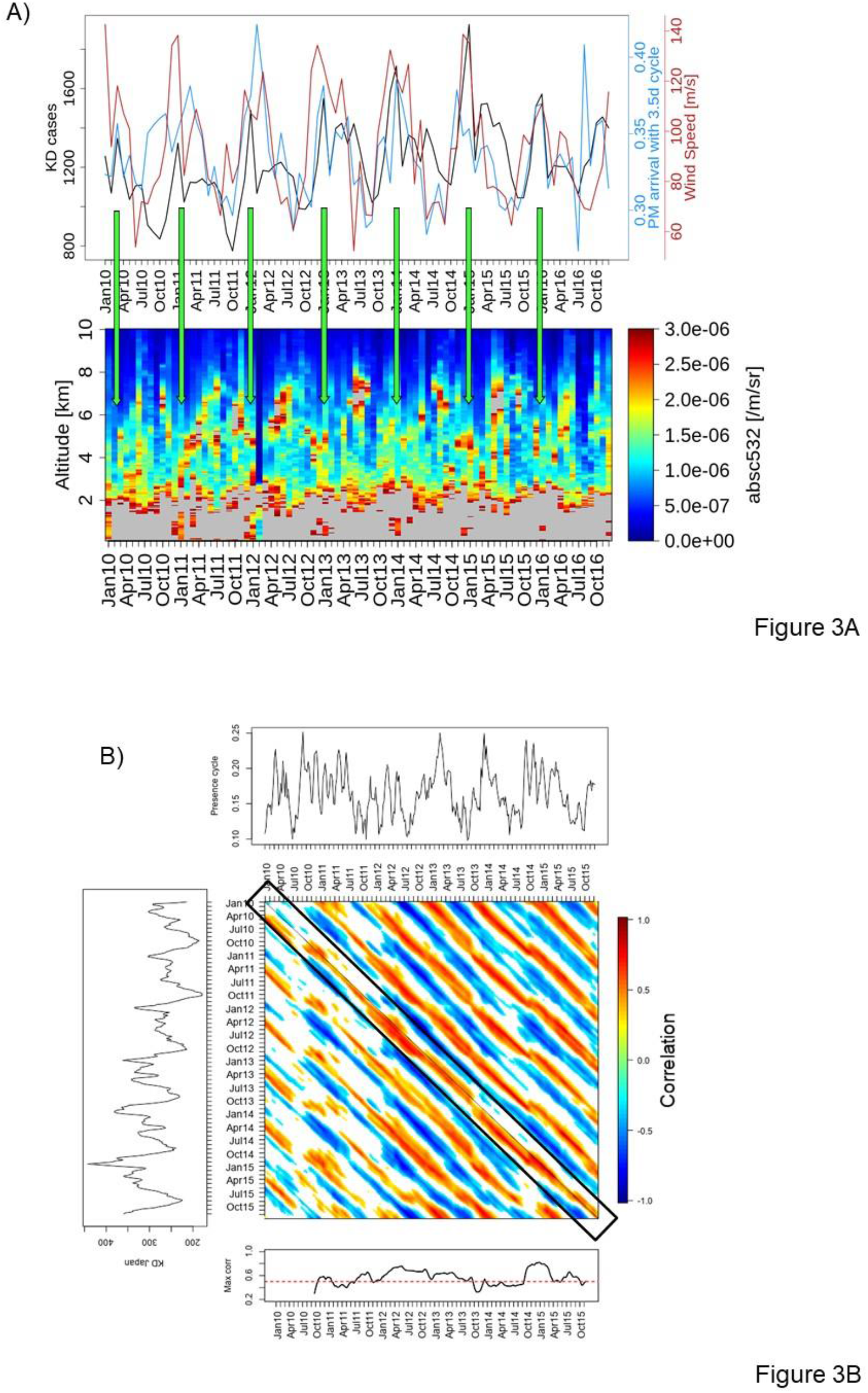
Association of particle-inferred abundance (absc532) arrival to Tokyo and KD cases in Japan. A) Temporal evolution of KD in Japan (black), Tokyo atmospheric PM_1_ arrival with SWC1 (3.5d) dynamics (absc532, blue; SWC-PM_1_) and wind speed(dark red); monthly integrated. Black box denotes surface air intrusions of windborne particles from high altitudes (circa 6km). These intrusions are displayed in the monthly averaged LIDAR profile for absc532. B) TW-SDC analysis between the WC1 series in absc532 LIDAR-inferred particle concentrations over Tokyo (top series) and KD weekly total cases in Japan (left series). Analyses in B) are performed over weekly data (week 1 to week 313 covering the total interval of time between January 2010 and December 2016). Window size (s) for B) is 52 weeks, corresponding to one year. Bars there denote Spearman correlation values and bottom panel shows average maximum correlations per week in a time lag going from 0 (phase) up to 15 weeks (with absc532 leading KD as in the black box).

Seasonality in these same variables for 2010–2016 was studied grouping monthly changes in the three aforementioned variables for Tokyo, Toyama and Tsukuba (Fig. 4B). These three locations are unique in that all have routine LIDAR instruments being operated from 2010–2016 (http://www-lidar.nies.go.jp/AD-Net/index.html), as well as long-term KD epidemiological records. KD in Fig. 4B groups all dates with KD maxima by month (red: 90% threshold, see Methods).

To expand on results above, we applied an SDC correlation analysis (48,49) in Fig. 3B between the same two variables during 2010–2016. A strong link between them appears that enhances towards the present, and denotes a leading role for airborne particles on KD (top series). This lead is clearly evident as strong positive associations (red dots) denote correlation values above +0.6 and up to +0.8 (*p< 0.01)* (see black box in lower panel) present at and below the main diagonal. Atmospheric PM over Tokyo was studied in relation to the WC up to 6km height and the direction of their movement traced (53). This way, downward direction in the air column of particles arriving at Tokyo (PM_1_ analog: blue line, see Fig. S5 and Methods for the PM_1_ size determination) (51) is shown in Fig. 3A (monthly) and Fig. 4A (weekly) for the entire 2010–2016 interval, together with wind speed (red line) and KD incidence (black line). Clear covariation is evident between PM_1_ and KD maxima occurring both also at times of maximum wind speed, and not with higher air column stability. These maxima correspond to air intrusions and downward motion over Tokyo as indicated by the black box in the monthly averaged LIDAR atmospheric profile. High(Low) seasonal concentration of aerosols exists in the surface due to strong(weak) downward winds typically occurring in winter(summer) since 2010 (Fig. 3A, 4A). Both monthly (Fig. 3A) and weekly (Fig. 4A) evolution in the three aforementioned variables further reinforce this strong co-variation. Predominance of the SWC1 at different heights clearly displays a downward movement as denoted also by SDC analyses, with larger amount of SWC1 variability decreasing as we move down the atmospheric profile. Despite causality cannot be directly inferred, results strongly point towards a consistent link among them all and to the involvement of tropospheric aerosols in the epidemiology of KD.

**Figure 4.**
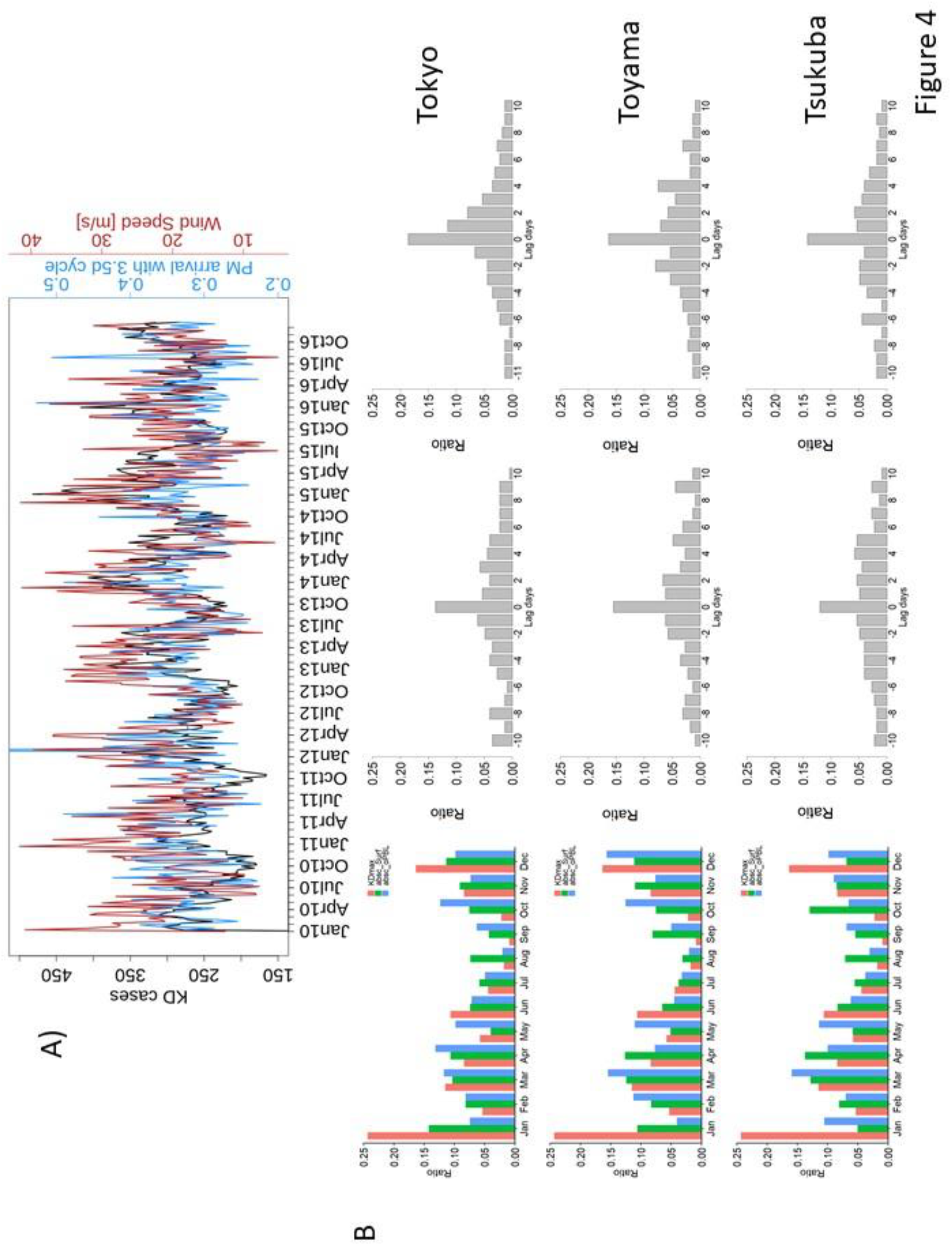
Coherence among LIDAR stations in seasonality, weekly association (Tokyo) and daily synchronicity of external PM arrival dynamics (WC1) with KD in Japan. A) Top panel denotes the weekly association between PM_1_ arrival in Tokyo with SWC1-PM_1_, blue) and KD in Japan (black). This vertical transport, from 4.3–6km towards the surface (120–600m), is once more backed by the wind speed (red) covariation(same methodology and analysis as Fig. 3A (top) in weekly scale). B) Co-evolution of KD and synchrony with the 3.5d cycle (SWC1) both over the PBL and in surface air over three LIDAR stations. Left) Seasonality in KD maxima (red) for the entire Japan and absc532 with SWC1 over the PBL (oPBL, blue) and at the surface (surf, green). Bars are normalized in respect to the total amount of maxima (KD), correlation values (absc532) per month. Middle) Histograms of the lag in days between absc532 respect to KD maxima oPBL, normalized by the total amount of dates from KD maxima. Right) Idem as Middle but at the surface. Rows indicate Tokyo (top), Toyama (middle) and Tsukuba (bottom).

Fully coherent seasonal changes exist also in SWC1-PM_1_ both over the PBL (oPBL, blue) and at the surface (surf, green) in the three locations. Daily delay between the disease and SWC-PM_1_ at both the surface (Fig. 4B, right column) and over the PBL (Fig. 4B, central column), supports their relation at a daily time scale since the lagged distribution is zero-centered (see also Fig. S5B). Dominance in all three locations occurs at zero lag, indicating there is no daily delay between occurrence of maximum in both atmospheric particles and KD incidence.

### Association of metal elements in PM1 particles and KD

To further explore the characteristics of particles potentially related to KD outbreaks, we used daily monitoring of PM_10_ (PM< 10µm) and their chemical analyses for Kumamoto (southwest of the Japanese archipelago) in spring 2011. Nearly 60 major and trace element concentrations (mostly metals) were determined from filter samples collected over 37 consecutive days (see 54 for a full description of the analytical methodology). Results demonstrated a remarkable covariation when compared to similar elements measured at 33 air quality monitoring stations in urban, suburban and rural areas operated by the Kumamoto Prefectural Government (see also http://taiki.pref.kumamoto.jp/kumamoto.taiki/index.htm). Fig 5A shows the stunning covariation between the total daily content of metals (Met_tot_, red line) and KD incidence (KD_03_, denoting sum of new cases that are diagnosed the same day or up to three days later, black line) in Kumamoto. Covariation of KD_03_ with most abundant individual elements is evident also throughout the 37 days of the monitoring campaign, with predominance of certain metals (e.g. Zn, Pb, Mn, Ba, Cu, As, Se, Bi and Tl) (Fig. S6). A linear regression fit was the best model relating metals in Fig. S6 (Se, Cu, Pb, Zn, Ba, Mn, Bi, As) and KD cases recorded in days 0 to 3 (Fig. S7 and Table SI, with lowest AIC: 43,018; r^2^: 0,727, p=5,08*10^−7^). Percentage of variance accounted for by metals in that specific atmospheric event rose up to over 50%, reaching values near 60% in the case of Zn (Fig. S7; Table S1). A linear regression model denotes how an increment of around 70 ng/m^3^ in the concentration of total metals is associated with a new KD case (Fig. S7). Further SSA analyses applied to the total amount of metals in Fig. 5A yielded clear separation of variability into four pairs of significant components, namely two in Fig. 5B having a 15d periodicity that quite stably varied as the oscillations reconstructed for KD_03_. Two other paired components in Fig. 5C recover exactly the SWC in KD, being the 15d cycle harmonic of the SWC. Strikingly, coarse particulate matter (denoted by PM_10_) and total carbon content (C_tot_) in Fig. 5B and 5C also display the same two main periods above, albeit they peak with slight delay in the later part of the survey and with a growing (convoluting) amplitude. Both C_tot_ and PM_10_ are not concurrent with neither Met_tot_ nor KD_03_ instead, whereas interestingly, Fig. 5B and 5C show how the evolving amplitudes of the reconstructed SWC components are exactly the same between metals (Met_tot_) and KD_03_ (often with even a one-day lead time).

**Figure 5.**
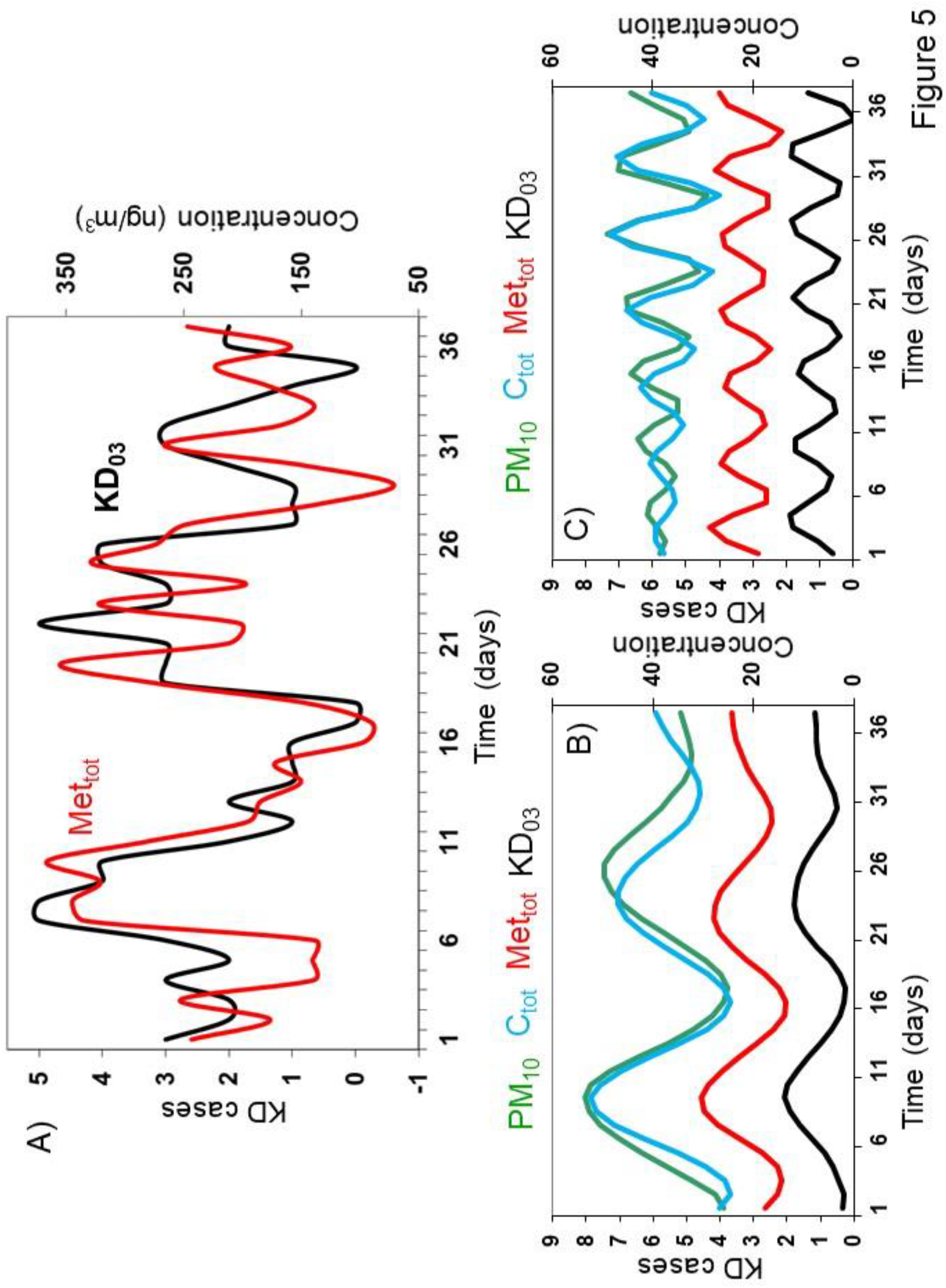
Relationship between air chemistry and new cases of KD during air intrusions at the Kumamoto prefecture (south of Japan) A) Co-evolution of Kawasaki disease increase in cases and total metal concentrations in a 37–days continuous surface air monitoring campaign at Kumamoto (Kyushu, see methods). KD cases are integrated over day 0 (arrival of air masses) and up to day+3 (KD_03_) to capture patients arriving from day zero to manifest the disease three days after (*8*). B) SSA reconstructed variability at the bi-weekly time scale (see methods and *19*) in the case of total metals (brown), total carbon (green), PM_10_ (red) and KD (black). C) Same than B) but for the weekly reconstructed components. Note consistencies and differences among phases and convoluting amplitudes of the reconstructed signatures in the different variables. Embedding dimension for the SSA analysis was always varied between 30<M< 50 to check for signal stability (*p< 0.05*). Concentration is in ng/m3 for MET_tot_ and in µg/m3 for Ct and PM_10_.

In the interval of roughly one month of daily sampling, different atmospheric situations were described that gave rise to the varying PM_10_ chemical composition described in Fig. 5 for metals and C_tot_. A first event around 17–27 March 2011 with NW winds feeding in transient diluted waves of aerosols from Asian mainland, originated temporary peaks of the fine particulate fraction (0.5µm– 2.5µm) and yielded spikes in several metals (54)(Fig. S6). Metals are enriched in varying degrees in the different size distributions and varying concentrations of Zn and Pb and to lesser degree also Mn, Ba, Se and Cu are evident with coherent PM_10_ and C_tot_ loads, denoting a mixture of dust and aerosol of small sizes (*not shown*). A mixture of dust and pollution aerosols from different parts of East Asia (e.g. stagnation over central China, Gobi Desert plume from the NW and agricultural debris) creates a complex air quality plume. Exposure to some of the main metals found (e.g. Zn, Cu and Pb) has shown high correlation with pulmonary inflammation by inhalation toxicology studies using animal models (55). The long- and short-term exposure to these hazardous air pollutants, individually or collectively, may cause respiratory (eye, nose, throat and sinus) irritation, allergic dermatitis and adverse cardiovascular effects (56). Exposure to abundant, highly oxidative, metal-rich pollution nanoparticles is a plausible route into cardiovascular pathogenesis (e.g. concentrations up to ∼22 billion magnetic nanoparticles/g of ventricular tissue were found in hearts) (57). Inhalation of air pollutants induces pulmonary oxidative stress and inflammation and presence of soluble transition metals in PM_x_ enhances the inflammatory responses via increased oxidative stress and the release of reactive oxygen species (58). Fine (PM_2.5_) and ultrafine particles (< 0.1μm) induce innate immune response via reactive oxygen species generation by transition metals and/or polyaromatic hydrocarbons (59). PM_x_ toxicity can be modified due to concurrent bioaerosols contribution or by their ability to modulate the oxidative potential of toxic chemicals present in PM_x_ (60). Therefore, they modulate both the innate and adaptive immune responses (61–64) in ways that can be similar to those observed in KD pathogenesis, although specific research along these lines is underway.

## Conclusions

Long-standing controversy on the ultimate causes of KD as a paradigm human vasculitis, should now consider that both physical and chemical nature of aerosols play a determinant role in its onset and epidemiology. Previous studies had essentially addressed variability at seasonal and longer time scales while discarded any detailed analyses on the day-to-day disease and aerosol changes. The discovery of a consistent sub-weekly signature in all the KD epidemiological records, at both the prefecture level and for entire Japan, present continuously since the early 1970s, sheds new light on the necessary concurrent factors leading to KD. This variability has long been confounded by a weekend bias in reporting but our results clearly show that the fundamental cycle of 3.5d does not match with a mere week-end effect. Instead, variability at this scale can only be linked to the anthropogenic effect on air quality with two weekly minima and maxima. These same patterns are effectively traced in air particles at all levels in the lower troposphere, being enhanced at times of air intrusions (and concurrent with KD maxima). Clear weekly cycles of anthropogenic origin, such as for NO_2_ and PM_x_ have been described by satellite measurements (e.g. GOME) (38,42). However, the fact that SWC components altogether account at most for around 20% of the overall KD variability, denotes that factors conducive to SWC are, however, not sufficient to determine alone a KD outcome. The former is in agreement with other studies where seasonal-to-interannual changes were studied as the main source of KD variability (15,18). For instance, trend in KD is possibly related to other processes distinct to the SWC here described, e.g. increases in cropland yield and cover (65). The Asian summer monsoon is known to effectively pump Asian pollutants to the upper troposphere and lower stratosphere, leading to enhanced aerosol formation that are then exported to the entire Northern Hemisphere (66), and tend to subside in Japan (67,68). However, although it is clear that KD enhancement is associated to external air intrusions, and temporary associations with aerosol’s metals can even account for over 50% of the total variability in this disease, we do not discard that a similar but less pronounced inflammatory effect can equally be exerted on KD susceptible children during low incidence days by local air pollution. It also remains to be investigated as other studies have suggested, whether embedded in aerosols and affected by their chemistry, other factors such as microbes or organic side-products, in genetically predisposed children, might also be playing a key role in KD pathology and in other similar vasculitis diseases.

## Data Availability

KD datasets were kindly provided by Prof. Y. Nakamura at Jichi Medical Hospital by means of Japan nationwide surveys. LIDAR data were kindly provided by Atsushi Shimizu and the AD-Net database at NIES, Tsukuba (http://www-lidar.nies.go.jp/AD-Net/index.html).

## Acknowledgements and Data access

KD datasets were kindly provided by Prof. Y. Nakamura at Jichi Medical Hospital by means of Japan nationwide surveys. LIDAR data were kindly provided by Atsushi Shimizu and the AD-Net database at NIES, Tsukuba (http://www-lidar.nies.go.jp/AD-Net/index.html). We would like to thank Jane Burns, Teresa Moreno, Xavier Querol, Dan Cayan, Josep-Anton Morguí, Atsushi Matsuki and Hiroshi Tanimoto for their helpful assistance and data sharing. This research was conducted with the financial support of the European Union’s Horizon 2020 research and innovation programme under Grant Agreement No.813545 at the ADAPT SFI Research Centre at Trinity College Dublin. We also acknowledge the Daniel Bravo Foundation for their financial support through a research grant to WINDBIOME, the HELICAL ITN EU Marie Curie and the KD parents’ association Asenkawa for both their economic and enthusiastic support.

## Author’s contributions

X.R. took the leading role in the conception and design of this study, A.N.-G. participated in the design, X.R., A.N.-G., S.B., J.C.B., T.K., A.M. and Y. N. participated in the acquisition of data, A.N.-G. and X.R. analyzed the data and all authors participated in the interpretation of data and discussion of results. X.R. drafted the article and all authors edited and approved the manuscript.

## Code accession

All codes are available on request to the corresponding author.

## Methods

### Datasets

Epidemiological data used in this study for Japan are total daily counts of KD patients admitted to hospitals for each of the 47 prefectures of Japan. To account for possible effects of different population sizes in cities, incidence was also calculated for comparison in the case of major Japanese cities from population censuses interpolated from 10 years national demographic records and only small differences emerged with results obtained for cases. KD cases were then weighed against the total pediatric population in that prefecture, with data covering the interval 1977–2016 (see KD case ascertainment protocol for Japan sites) (*8, 15*). The Japanese KD time series is derived from the biannual to 24 separate questionnaire surveys of Japanese hospitals for the period 1970–2016, national surveillance of all hospitals in Japan with more than 100 beds. Data corresponds to 24 separate questionnaire surveys of Japanese hospitals for the period 1970–2016, providing the most comprehensive record of KD cases in the world. Patient date of hospitalization and prefecture of residence are recorded for each subject in the time series. Time series are formed by KD patients, the date being that of hospital admission recorded for all subjects. For subjects with multiple admissions, only the first hospitalization date was used. To provide uniform assessment among sites, the date of admission was used as a surrogate for the date of fever onset, even though for comparison with other studies, the average day of KD diagnosis is normally centered on the 5th day of fever. However, analyses were reproduced with calculated dates of fever onset and results did not differ (*p< 0.001*, one–sided t-test). Approved written consent was obtained from all participants. Human population epidemiology studies were approved by the Bioethics Committee for Epidemiologic Research, Jichi Medical University.

NCEP/NCAR Reanalysis 1 project wind data (u-wind and v-wind components) were used from 1970 to 2016 (*68*). Wind speed (ws) was calculated as: ws: u = ws * cos(θ) and, v = ws * sin(θ). Data (m/s) was extracted from the nearest grid cell to Tokyo (LON = 138.75, LAT = 37.14). City population Japan was obtained from http://poblacion.population.city/japon/.

LIDAR-inferred particle variable is obtained from pre-processing the AD-Net LIDAR (http://www-lidar.nies.go.jp/AD-Net/ncdf/) NetCDF files with an R script, using the package *ncdf4*. These files provide data from the following lidar parameters:

i. 532nm attenuated backscatter coefficient [/m/sr] (absc532)
ii. 1064nm attenuated backscatter coefficient [/m/sr] (absc1064)
iii. 532nm volume depolarization ratio (dep532)
iv. 532nm aerosol extinction coefficient [/m] (ext532)
v. 532nm dust extinction coefficient [/m] (extd532)
vi. 532nm spherical particle extinction coefficient [/m] (exts532)
vii. 532nm aerosol depolarization ratio (adep532) viii. Estimated mixing layer height [m] (hPBL)

Time and height are the two dimensions of the LIDAR parameters making them spatiotemporal variables. Time resolution is 15 min, with 96 profiles per day and height integrations every 30m from the surface up to 15km. These values are accumulated and averaged with even higher resolution, in order to increase the signal-to-noise ratio as to five range bins for height and three for time. Data is received every hour and it is converted to optical parameters through a processing program. Corrected data results neglect values below 120m since the accuracy cannot be computed due to limitations in the geometrical overlap correction factor.

From the seven air particle related parameters (all except hPBL) listed previously, there are three with low level data preparation: dep532, absc532 and absc1064; and four for classification of scatters and estimation of optical parameters: ext532, extd532, exts532 and adep532 (*46*). These last parameters are calculated from the low-level data preparation parameters. Therefore, we selected absc532 as the aerosol parameter because it can detect smaller particles than absc1064, and it retrieves information about the abundance and size of air particles, while dep532 its sphericity.

In an attempt to enhance the signal-to-noise ratio, we integrated data by atmospheric layers and this way we gathered information of dynamics above and below the PBL. To this end, we selected the surface level (120–600m), a height right above the PBL (oPBL, 1400–2500m) and two higher layers to investigate the potential external origin of aerosols. The latter were layers integrating 2800–4000m and 4300–6000m heights. This distinction of layers helps understanding the origin of the time series dynamics since Tokyo particles are proven to follow a downwind transport (Fig. 3 and 4). Therefore, a vertical transport is categorized as downwind transport if the dynamics on the higher atmospheric layers anticipate that at the surface. We inferred this PM_x_ transport is most often external since the origin is from above the PBL.

### Methods

We used the SDC analysis (*48–50*) to study the local transitory nature of patterns in every series studied as well as the local correlations existent between different variables. The one-way SDC (OW-SDC) technique is an univariate method that computes non-parametric Spearman rank correlations with itself using rcorr (https://www.rdocumentation.org/packages/Hmisc/versions/4.2-0/topics/rcorr), at a variety of window sizes (s). A correlation value is then obtained at a given significance level (*p < 0.05* or *0.01*, as indicated). SDC technique is sensitive to s; and therefore, different s above and below the period of interest are checked to assess for consistency in the results. However, despite being an optimal method for short and noisy time series, the resolution it yields cannot capture all the strength of the 3.5d cycle from just daily data as in the case of KD. That is because the correlation function needs at least 4 values as a feasible window size, s≥4 units. This limitation has been proved by analyzing synthetic time series built as x(t) = A*cos(2*π*F_c_*t), where F_c_ is the frequency, here 1/3.5 [days^—1^]; where A would be the amplitude of the oscillation which is set to 1, regardless of its fit with the peaks since the main interest is in phases. The OW-SDC is not able to identify the 3.5d cycle, retrieve the positive off diagonal at 3.5d, when the window size was at its minimum of 4d for the synthetic time series (not shown). Instead, it was able to properly extract this period and the harmonic at 2.33d at larger s (namely at s = 10d, such as in Fig.S1 and S2; albeit with lower correlation values due to this limitation). Coherently, the 7d period, a harmonic of the 3.5d (see main text and Fig. 1 and Fig. S1 and S2), was clearly visible and could be used as a marker for the 3.5d cycle since s = 7d. This assumption is also validated by the SSA analysis which attributes the half-weekly cycle as the main period, with the weekly cycle as a harmonic (Fig.1). Therefore, correlations for every aggregation time series in Fig. 2B (top) were extracted from the OW-SDC correlation matrix with s = 7d and considering also a lag of 6–8 units below the diagonal. The three off-diagonals are reduced to one by computing the mean of the positive correlation values per day, resulting in a daily correlation time series. The yearly count of daily positive correlations retrieves the evolution of the 3.5d cycle from its 7d harmonic.

An eigendecomposition analysis applied to the data covariance matrix of daily KD time series in Japan and San Diego was used to partition signal contributions by frequency, with the aid of adaptive nonparametric functions (*69*). An embedding dimension or order of the decomposition of 40 was selected, as it allows a proper characterization of those signals with period larger than the annual cycle, but much larger values (e.g. M = 200) proved totally stable in terms of both decomposition and reconstructed components (*2*0). The significance level of the spectral decomposition was computed using a bootstrap method (*70*). The reference probability distribution function was locally computed by means of 10000 white-noise time series, which were then tested against an autoregressive process of order 1 having the same mean, standard deviation, and temporal one-lag autocorrelation value as the interannual long-term KD time series (find further details in *18*). An autoregressive spectrum (AR) Spectrum with three different algorithms comparison with an order field ranging between values of 20 and 50 was computed on the original daily KD dataset. The AR high-order model is an optimal estimator of contributing frequencies as the frequencies depend only on the roots of the fitted polynomial. Each spectrum is also calculated independently and intercompared. We used the data singular value decomposition (SVD) algorithm for its robustness, although the FB (forward-backward), Fwd (forward), and Bwd (backward) prediction variants were also calculated for comparison. A 50% change in the signal subspace was also imposed to test for stability in the spectral signatures and no change in resulting frequencies occurred. As the data is not noise-free data, high orders were selected, well above the minimum order number needed (e.g. at least double the number of expected sinusoids in the spectrum). Runge-Kutta procedure was used to integrate the spectrum adaptively and to achieve a proper signal-to-noise discrimination.

### LIDAR analyses and PM arrival calculations

LIDAR-inferred particle dynamics are identified by its presence magnitude, which tracks a specific cycle for unit of time. SDC methodology (*48–50*) identifies these transient dynamics in the hourly scale for absc532. For WC1, we selected the maximum correlation values per hour from lags 76–92h (84±8h) computed by an OW-SDC analysis at s = 84h. This tolerance of ±8h is due to the nature of data, which produces fluctuations on the 84h cycle. These correlation values are then integrated in the temporal scale, providing a representation of its presence for each unit of time. Therefore, presence means more hours with the transient cycle favoring higher correlation values per unit of time. Presence of 3.5d period in absc532 is displayed in Fig. 3A (bottom) and 4A (top panel series) by averaging among the 4 atmospheric layers previously described and integrating by month. Both variables in Fig. 3A are smoothed by a local polynomial regression (https://www.rdocumentation.org/packages/stats/versions/3.6.1/topics/loess) with a loess method, with a second-degree polynomial and 0.1 smoothing degree parameter.

The PM arrival with 3.5d cycle variable from Figure 3A (top, blue), 3B and 4A (top) has been computed with a two-way SDC (TW-SDC) analysis, a bivariate method. The TW-SDC technique displays the transitory forcing of the atmospheric layers dynamics to the surface. In this case, the driving or forcing is from 4300–6000m towards 120–600m for the abs532 hourly time series. In the TW-SDC correlation matrix, the sector below the main diagonal represents the delay between both time series. In this case, the window size is set to 84h (3.5d) to discern the presence and interactions of WC1. The off-diagonals from 0–24h are extracted and the mean value per hour is computed, since downwind transport may occur within the same day. Therefore, we have now an hourly time series attributed to the PM arrival with the presence of the WC1. In Fig. 3A (top, blue) and 4A (top), we averaged the correlations values monthly and weekly (respectively) to study its relationship with the integrated wind speed and KD Japan cases. We believed that the mean is a more realistic parameter than the integration, due to the nature of the data and the pre-processing needed for the PM arrival with 3.5d cycle variable.

### Maximum Analysis

To further explore the relationship between KD and PM_x_, we developed a technique called ‘maximum analysis’ for both variables during the 2010–2016 period. First, we grouped KD Japan cases time series by year and computed the 90% of the values to gather the maxima and additionally remove the concomitant influence of the trend. Accordingly, we used the emp.hpd formula (https://rdrr.io/cran/TeachingDemos/src/R/hpd.R), from muscles of caffeine-supplemented for asymmetric densities. We sorted the maxima dates by month and normalized them by computing the ratio to the total number of maxima selected (Fig. 4B). This type of visualization proves useful to visualize the seasonality of the KD maxima. Second, we applied the methodology to track the presence of WC1 with OW-SDC, at both the surface and over the PBL (1400–2500m), for the absc532 hourly time series. Afterwards, the hourly timedates from the correlation values above +0.5 are gathered and sorted by month. These monthly counts are normalized by the total amount of dates used to provide a relative value comparable to the KD maxima (Fig. 4B left column).

Finally, we also computed the lag between KD maxima and the presence of the WC1 in a PM_x_ variable above and below the PBL (Fig. 4B central and right column). These bar plots display a clear distribution centered at 0, which synchronically associates the different variables. Therefore, KD evolution in Japan (2010–2016) association with the LIDAR-inferred particle dynamics (WC1) is coherent among LIDAR stations in seasonality, weekly association (Tokyo) and daily synchronicity.

## Sub-weekly cycle uncovers the hidden link of atmospheric pollution to Kawasaki Disease

**Fig.S1:**
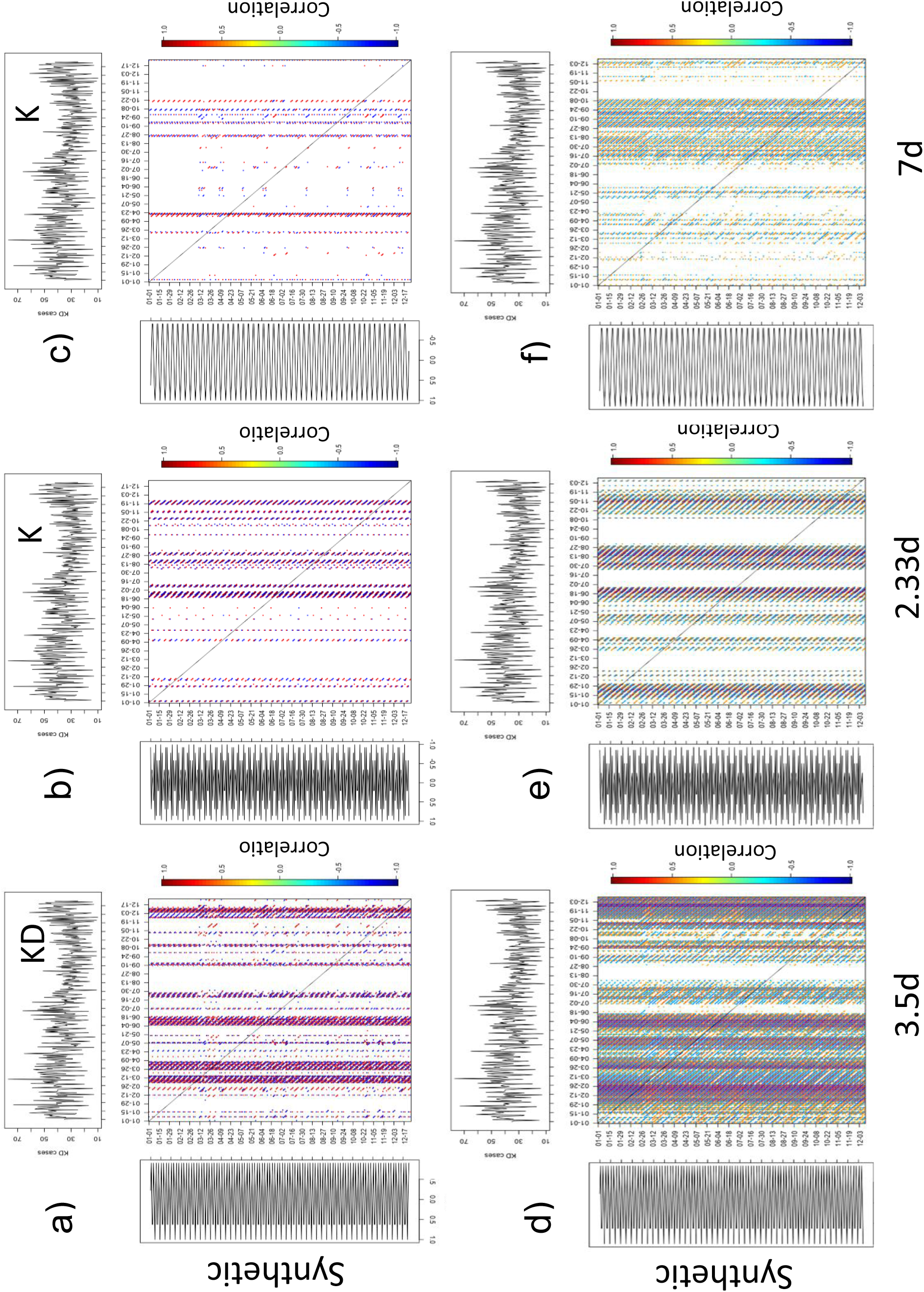
Total contribution of each pair of RCs to the overall WC variability in year 2010. TW-SDC analyses between KD in Tokyo (top series) and a synthetic series having the pure WC1 (3.5d), WC2 (2.33d) and WC3 (7d) were computed for 2010 (e.g. very similar patterns can be found for all years). Two different window sizes are used to test for stability in the results (top, s = 6d and bottom, s = 22d). WC2 is shown in middle plots (b and e). Note the much larger contribution of WC1 RCs (panels a and d) than WC3 (panels c and f) both in their respective appearances and in the magnitude of correlations, as well as the presence of the WC3 in the low KD season. WC2 is shown in middle plots(b and e). Patterns displayed are significant at least at the *p< 0.05* level (see Methods below for construction of the synthetic series).

**Fig.S2:**
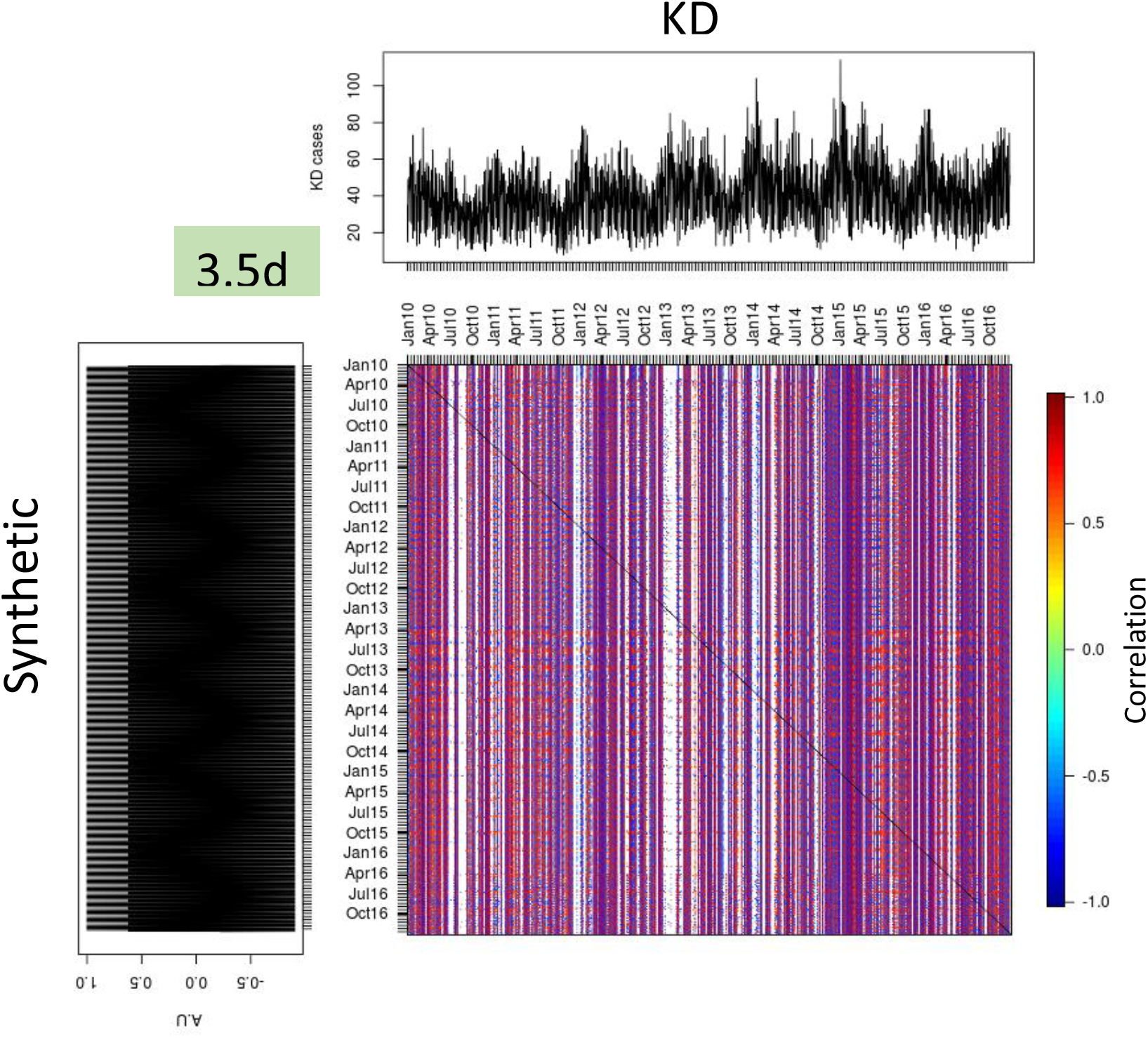
Total contribution of WC1 to the overall WC variability in the interval 2010-2016. Results of a TW-SDC analysis computed with the observed KD time series aggregated for entire Japan and a synthetic time series with a fundamental 3.5 days’ period. A significance threshold of 95% was imposed for correlations shown. Same results also hold for Tokyo and the nine aggregated prefectures in Fig. 2B.

**Fig.S3:**
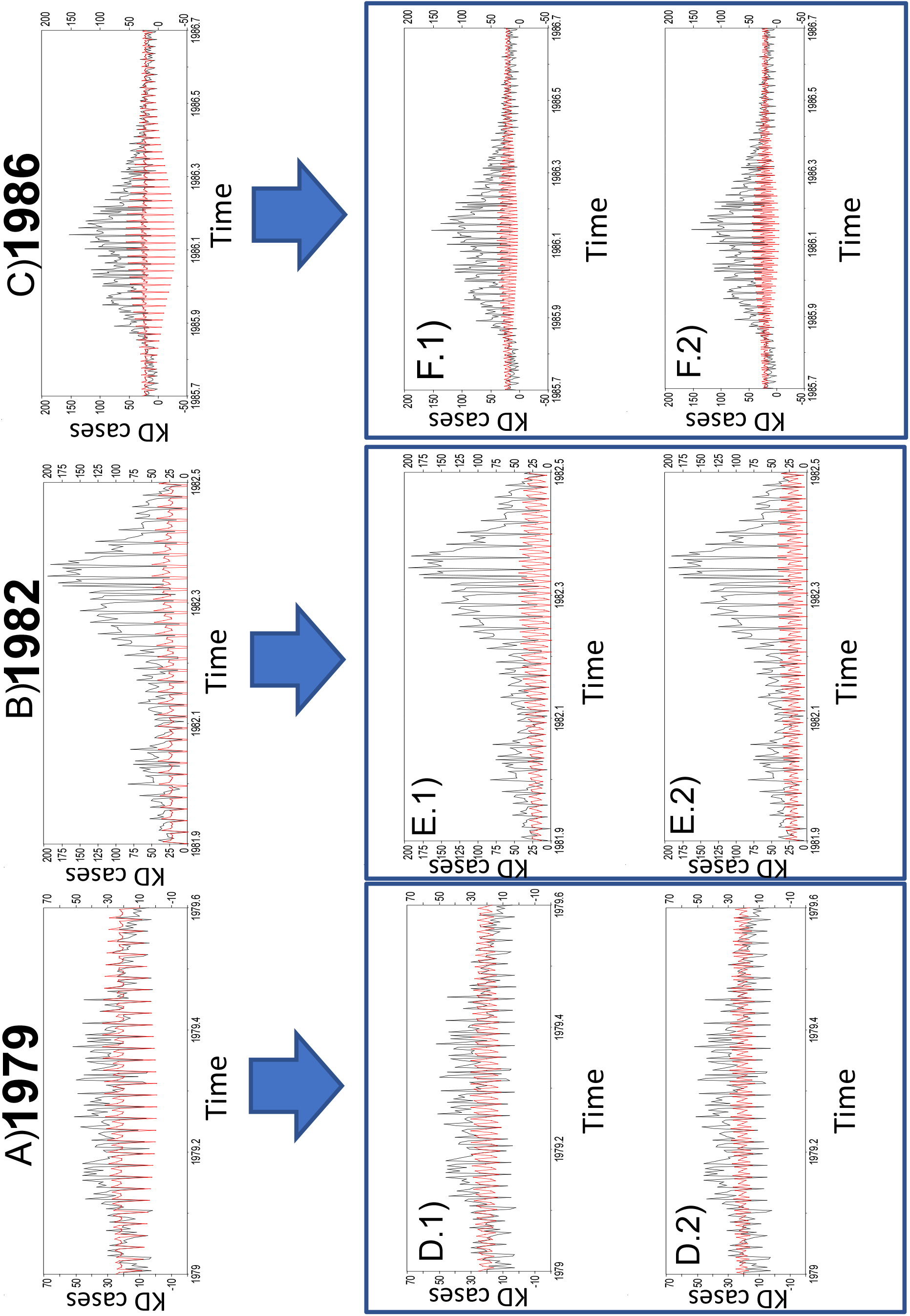
Reconstructed WCs in the three main KD epidemics (1979, 1982 and 1986) in Japan. For each epidemic, top panels show the contribution in each epidemic of the entire WC components. Bottom panels depict the two separate RCs in each year, namely WC1 (3.5d) in D.1, E.1 and F.1; WC2 (2.33d) in D.2, E.2 and F.2. Data shown is aggregated for all Japan.

**Fig.S4:**
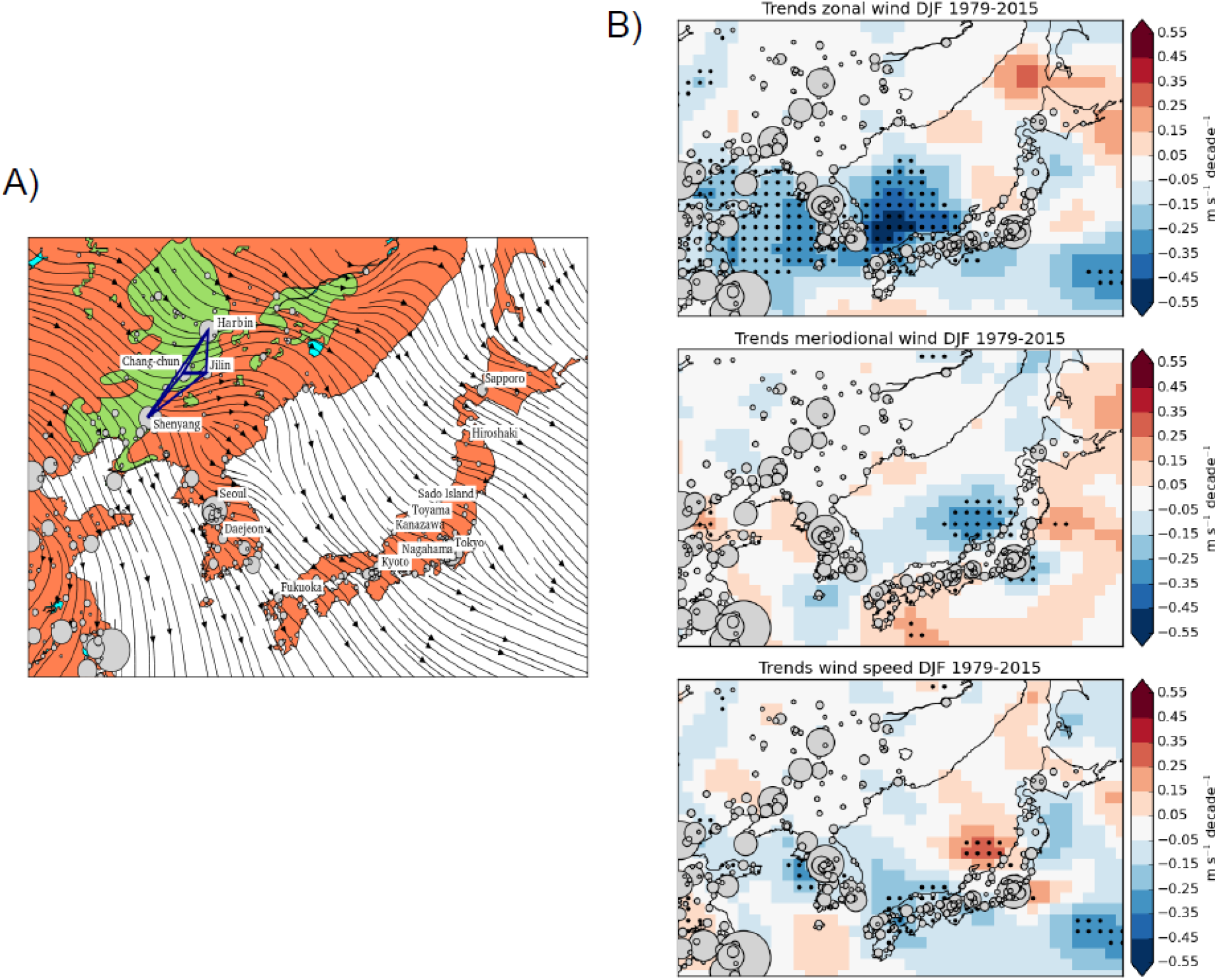
A) Gradient of mean winter (DJF) regional wind stress for the interval 1975-2015 in northeast Asia. Cities larger than 100000 people are shown (gray bubbles). For the highlighted cities in Japan, 15 years of LIDAR atmospheric column particle composition are available. The area of intensive cereal croplands is also shown (green) together with the area main cities. B) Trends (m*s^−1^/decade) in winter (DJF) zonal, meridional and wind speed in the interval 1979–2015. Dotted areas denote 95% significance levels.

**Fig.S5:**
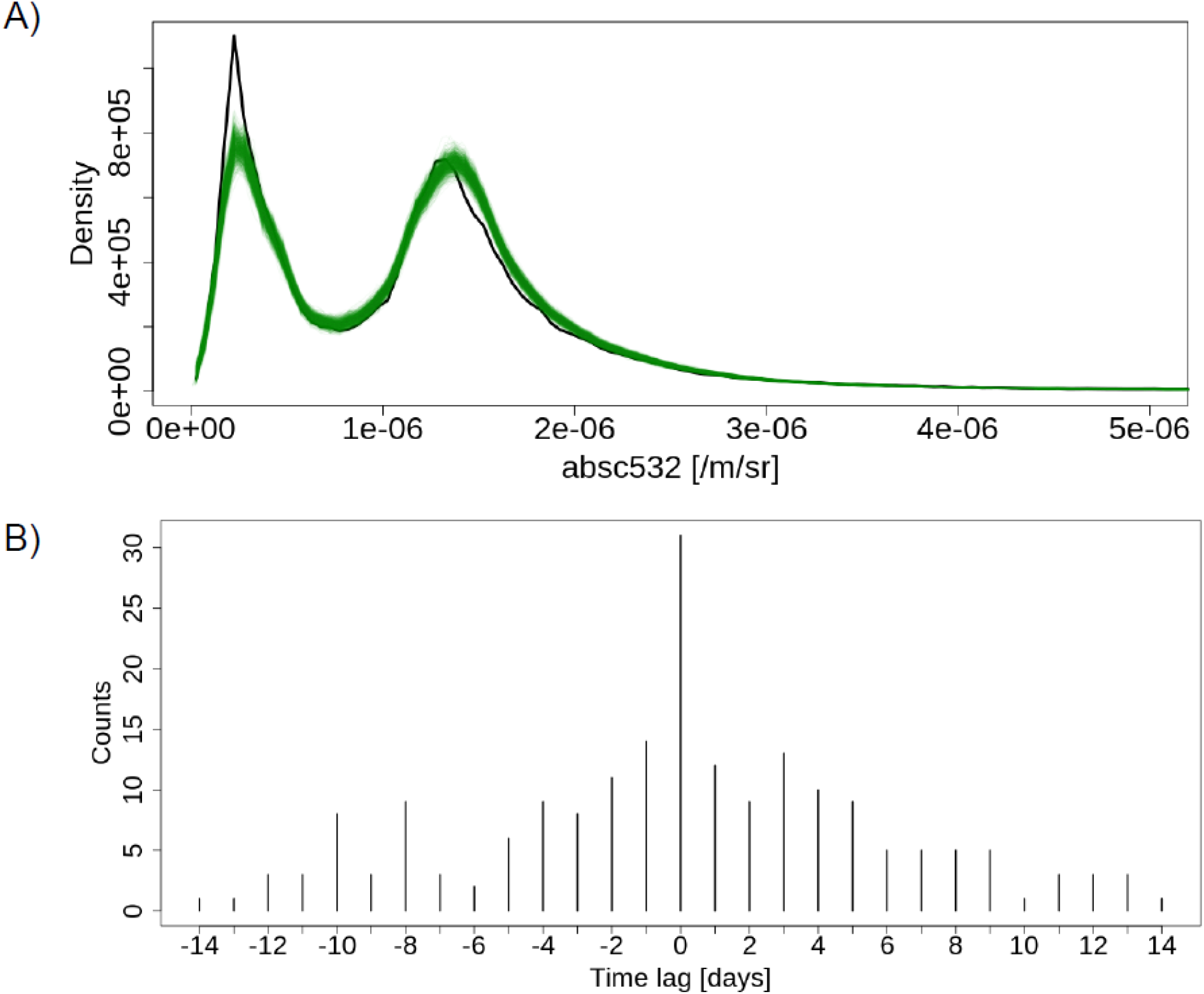
Particle-size distribution in the air column for Tokyo during 2010-2016. A)Observed distribution when WC1 is present (black) vs 1000 random distributions (green) in absc532 LIDAR measurements over the PBL (oPBL) in heights between 1400m and 2500m. B) Same as in Fig 4B (central column) but for total counts instead of ratios.

**Fig.S6:**
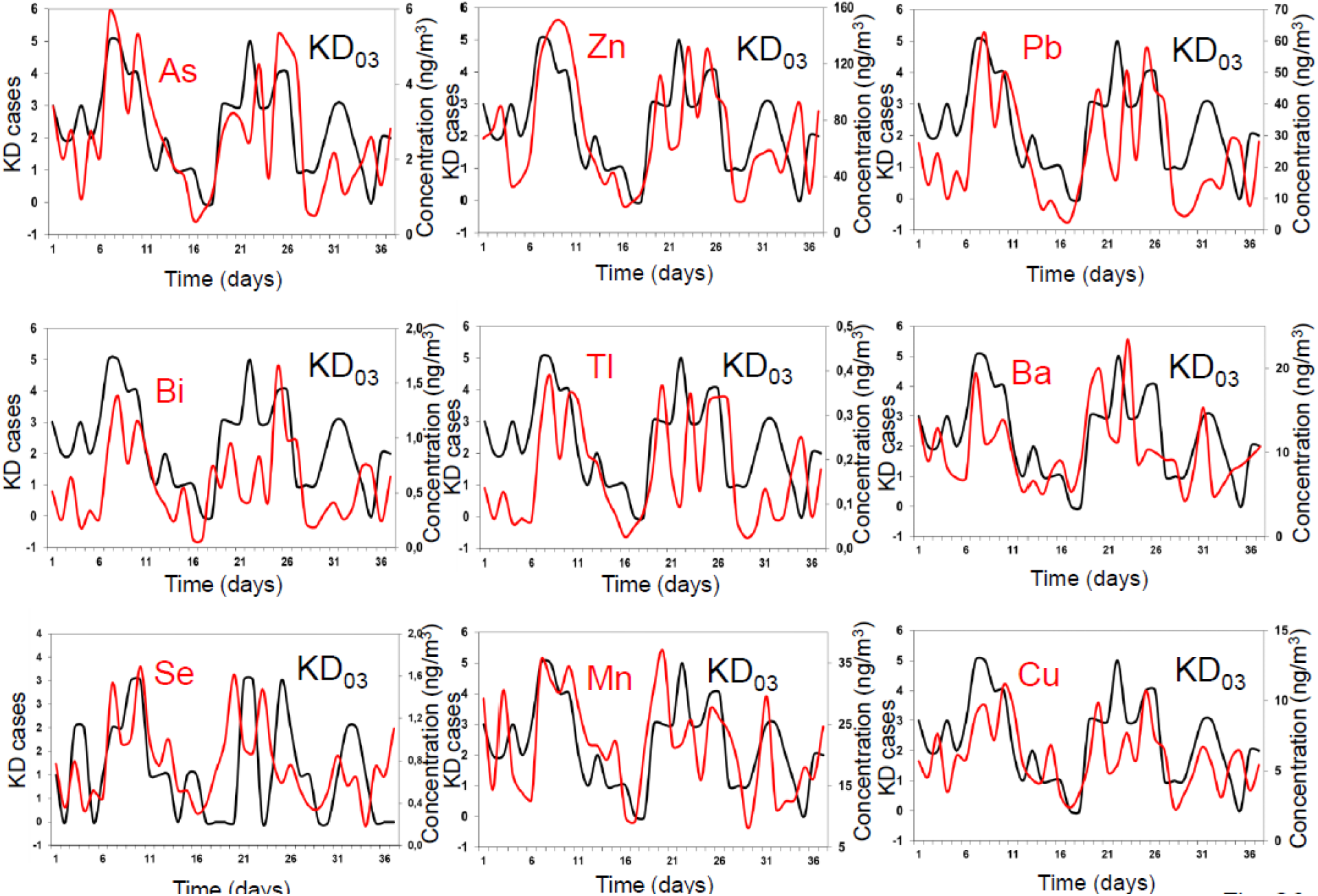
Same as Fig. 4A but for the most abundant chemical elements. (panels displayed are for elements related to KD disease with at least r_xy_>+0.6; *p< 0.05*).

**Fig.S7:**
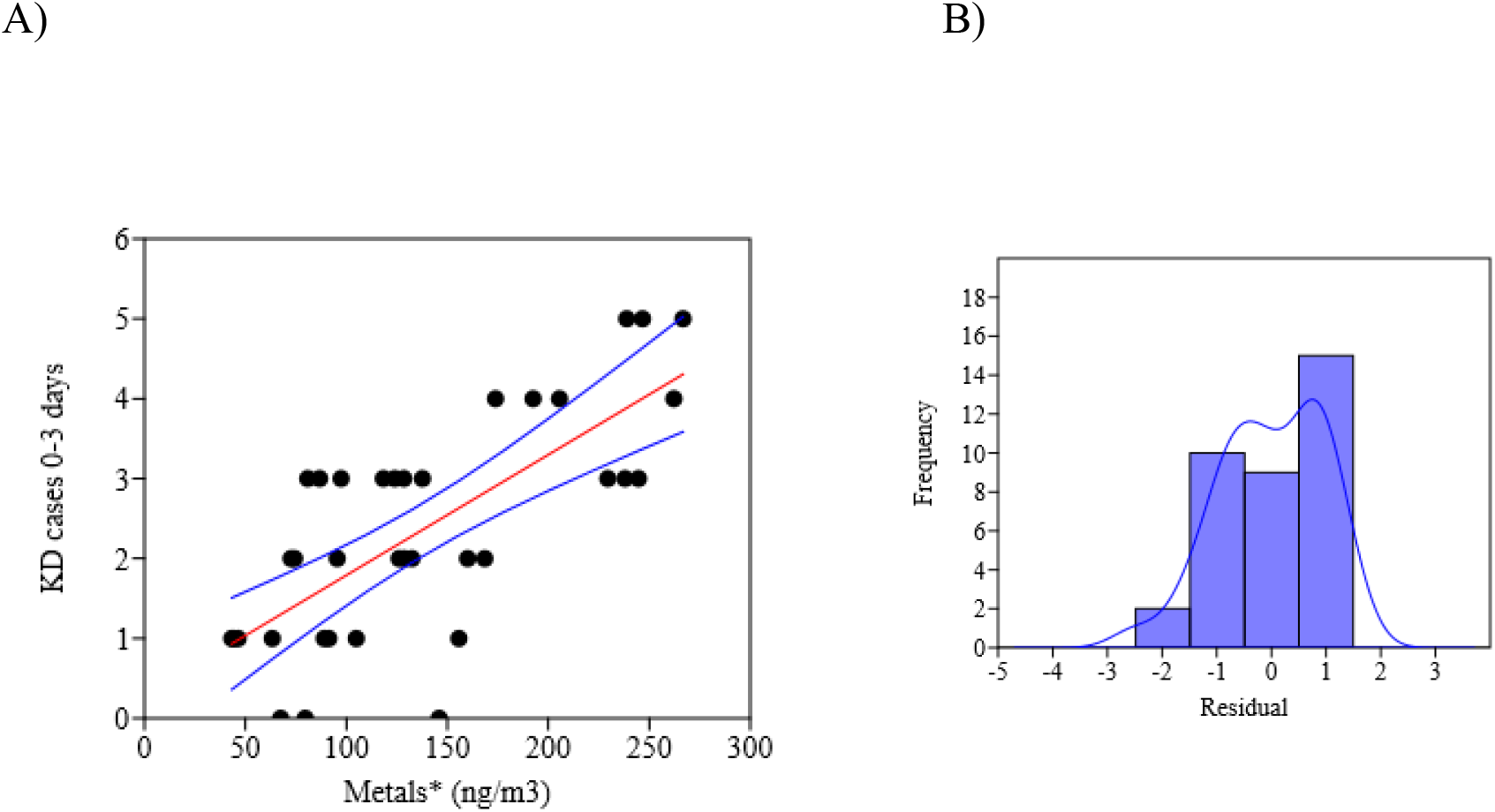
A) Ordinary Least Squares Regression between Metals in Extended Data Fig. 6 (Metals* (ng/m3) state for the sum of Se, Cu, Pb, Zn, Ba, Mn, Bi and As) and KD cases integrated for days 0 to 3 after air arrival (KD cases 0–3 days). B) Kernel density plot of the residuals showing a bimodal distribution. 95% bootstrapped confidence intervals highlighted as blue lines (N=1999). Correlation of 0,72718 (p=5,0794*10^−07^) accounts locally for over 50% of total variance in KD and denotes how an increment of around 70 ng/m^3^ in the concentration of total metals is associated with a new KD case. A linear regression fit was the best model relating metals in Fig. S6 (Se, Cu, Pb, Zn, Ba, Mn, Bi, As) and KD cases in days 0 to 3 (lowest AIC: 43,018; r2: 0,727 p=5,08*10^−7^). Fit is shown below with 95% bootstrapped confidence intervals (N=1999). Similar models applied to total metals yielded lower correlation values and higher AIC (45,369).

**Table S1.** Correlation values for all metals and KD03 in Fig. S6 (% variance also displayed for highest values).

| **Variables** | **Correlation** | **p-value** | **lag** |
| --- | --- | --- | --- |
| **Zn-KD03** | +0.768 (59.03%) | $4.487 \cdot 10^{-8}$ | 1 |
| **Pb-KD03** | +0.7025 | $1.7887 \cdot 10^{-6}$ | 1 |
| **As-KD03** | +0.69373 | $2.7154 \cdot 10^{-6}$ | 1 |
| **Tl-KD03** | +0.63763 | $2.881 \cdot 10^{-5}$ | 1 |
| **Cu-KD03** | +0.62481 | $4.634 \cdot 10^{-5}$ | 1 |
| **Mn-KD03** | +0.5659 | 0.00026118 | 0 |
| **Bi-KD03** | +0.55073 | 0.00050049 | 1 |
| **Se-KD03** | +0.54085 | 0.000548 | 1 |
| **Ba-KD03** | +0.5193 | 0.00098943 | 0 |
| **Met\*-KD03** | 0,727 (52.85%) | $5,08 \cdot 10^{-7}$ | 0 |
| **Met-KD** | +0,6492 | $1.38 \cdot 10^{-5}$ | 0 |

